# A multilevel approach to individual and organizational predictors of stress and fatigue among healthcare workers of a university hospital: A longitudinal study

**DOI:** 10.1101/2022.01.24.22269481

**Authors:** Oumou Salama Daouda, René Sosata Bun, Karim Ait Bouziad, Katiuska Miliani, Anastasia Essa-Eworo, Florence Espinasse, Delphine Seytre, Anne Casetta, Simone Nérome, Adelaide Nascimento, Pascal Astagneau, Laura Temime, Mounia N Hocine

## Abstract

**Objective:** Healthcare workers are at high risk of experiencing stress and fatigue due to the demands of their work within hospitals. Improving their physical and mental health and in turn, the quality and safety of care, requires considering factors at both individual and organizational levels. Using a multi-center prospective cohort, this study aims to identify the individual and organizational predictors of stress and fatigue of healthcare workers in several wards from university hospitals.

**Methods:** Our cohort consist of 695 healthcare workers from 32 hospital wards drawn at random within four volunteer hospital centers in Paris-area. Three-level longitudinal analyses, accounting for repeated measures (level 1) across participants (level 2) nested within wards (level 3) and adjusted for relevant fixed and time varying confounders were performed.

**Results:** At baseline, the sample was composed by 384 registered nurses, 300 auxiliary nurses and 11 midwives. According to the 3-level longitudinal models, some predictors were found in common for both stress and fatigue (low support from the hierarchy, low safety culture, overcommitment at work, presenteeism while sick…). However, specific predictors for high level of stress (negative life events, low support from the colleagues and high frequency of break cancellation) and fatigue (commuting duration, frequent use of interim staff in the ward…) were also found.

**Conclusion:** Our results may help identify at-risk healthcare workers and wards, where interventions to reduce stress and fatigue should be focused. These interventions could include manager training to favor better staff support and overall safety culture of healthcare workers.

**What is already known about this subject?:**

- Healthcare workers have high levels of perceived stress and fatigue, particularly in medical fields highly exposed to infectious risks.
- High occupational stress and fatigue can negatively affect healthcare workers behaviors in terms of absenteeism, and ultimately intention to leave as well as quality of care.
- Individual and organizational differences contribute to different perceptions and consequences of occupational stress and fatigue in healthcare workers.

**What are the new findings?:**

- The ward-level environment significantly influences the stress and fatigue of healthcare workers, in addition to individual factors and time variations.
- Hierarchy providing low support and with low safety culture, work overinvestment, presenteeism while sick, and working in smaller wards were identified as predictors of both high stress and fatigue of healthcare workers.
- Negative life events (whether personal or professional), low support from the colleagues and high frequency of break cancellation are specific predictors of high level of stress. While commuting duration, frequent use of interim staff and working in a medical ward were associated with high level of fatigue.

**How might this impact on policy or clinical practice in the foreseeable future?:**

- In this study, we can identify some areas for improvement to better prevent stress and fatigue for healthcare workers. High stress and fatigue can be reduced through mutual and specific organizational intervention strategies.

## INTRODUCTION

There is a growing research interest about stress in healthcare workers, as the prevalence of nurses affected by negative mental states is high [1,2]. This was underlined by a recent meta-analysis, included 45 539 nurses worldwide in 49 countries across multiple specialties, that estimated 11.2% prevalence of burnout among global nurses [3]. Moreover, the current COVID-19 pandemic has exacerbated stress, depression, and anxiety in front-line healthcare workers, as underlined by another meta-analysis [4].

In consequence, healthcare workers are likely to develop chronic health problems and stress-related illnesses which can result in frequent staff sickness and absenteeism [5,6]. Stress and fatigue in the workplace negatively impact productivity and absenteeism [6], and may result in non-optimal quality of care for patients in healthcare settings [7,8]. For instance in 2020, the average absenteeism rate in French public hospitals was estimated 9.5% among a representative sample of 300 hospitals [9].

Among healthcare workers, stress and fatigue both have multifactorial etiology and are different in nature. In fact, they cannot be explained just by the classic unicausal model of occupational illnesses but rather by the model of “work-related illnesses” [10]. Both organizational and personal factors are involved in healthcare workers’ occupational stress [11–13]. The latter are also exposed to classic and emergent psychosocial work factors and mental health [14].

While the occupational predictors of both stress and fatigue in nurses have been explored cross-sectionally before, for instance by Jones et al. in the French context [15], very few longitudinal studies are available on this subject [16,17]. In addition, there is still a lack of literature regarding the trajectory of stress and fatigue among healthcare workers that accounts for the specificities of the wards and hospitals they work in, in terms of structure and organization.

Here, using 1-year longitudinal data collected in 32 French hospital wards, we aim to determine the real-time associations between perceived stress and fatigue of healthcare workers, and individual and organizational-level factors.

## MATERIAL AND METHODS

### Study design and participants

#### Study Design and data collection

We designed a multicenter study on the individual and organizational predictors of stress and fatigue and infectious risk among healthcare workers at the Hospitals of Paris, the STRIPPS study [18]. The study was conducted between February 2018 and July 2019 and data were collected on midwives, registered and auxiliary nurses. Healthcare workers were recruited from 4 voluntary French University general care hospitals. Eight wards per participating hospital were drawn at random wards employing at least 30 healthcare workers.

Data were collected longitudinally every 4 months during one year by two different interviewers for all included participants as follows: t0, corresponding to the first collection during the healthcare worker inclusion visit; t1, t2, t3, corresponding to follow-up visits at 4 months (t0 + 4 months), 8 months (t0 + 8 months) and 12 months (t0 + 12 months). For the first data collection (t0), dates and times of visits by an interviewer were drawn randomly for each participating ward. For later data collections, individual appointments were made with each included healthcare worker. Data were collected by the interviewers through administered questionnaires at both levels (ward level and individual level).

#### Individual-level variables

- At the individual level, the following characteristics were collected:
- Part 1 included general and occupational characteristics such as age, sex, professional status, contractual situation, length of employment at the institution (years), and daily working hours.
- Part 2 consisted of the French version of the hospital survey on patient safety culture [19]
- Part 3 consisted of the French version of the Job Content Questionnaire (JCQ) [20] to measure support from hierarchy and colleagues
- Part 4 included general questions about work organization such as time schedule, nightshift, extra hours, mealtimes, and rest periods.
- Part 5 consisted of the Effort-Reward Imbalance at work questionnaire (ERI) to measure work overcommitment [21]

#### Ward-level variables

At the ward level, the following characteristics were collected: specialty, number of beds, proportion of double rooms, frequency of tasks performed outside the ward, healthcare worker to patient ratio, and use of external healthcare service providers (i.e., interim staff).

#### Outcome Measurements

Two primary outcomes were considered:

1. perceived stress, assessed with the Perceived Stress Scale 10-item scale (PSS-10). The PSS-10 score ranges from 0 to 40 (from very low to very high perceived stress). The PSS-10 questionnaire was developed by Cohen and al. [22] and validated in French [23,24].
2. fatigue, assessed with the Pichot fatigue scale. The Pichot score ranges from 0 to 32 (from very low to very high fatigue) [25].

### Data analysis

#### Missing data imputation

For missing data, multiple imputation was performed on validated questionnaire items only (JCQ, PSS-10, Pichot, and ERI questionnaires). Data were imputed using multiple imputation, using the R *mice* package [26]. The *mice* package allows to perform imputation of continuous and categorical variables in a context of multilevel and longitudinal data. Indeed, the multilevel structure of the longitudinal data need to be considered in the imputation model. All questionnaire items with missing data were imputed using joint modeling and chained equations [27] considering participants as clusters. For all questionnaire items included in the imputation model, missing data were assumed to be missing at random.

#### Statistical analysis and modeling

We conducted analyses to identify factors associated with the stress and fatigue levels of participating healthcare workers. First, in order to validate the use of a 3-level longitudinal model, we built two unconditional models (i.e., null models, with no independent variables) with two levels (i.e., time and individual levels) and three levels (i.e., time, individual and ward levels), for each outcome. In fact, before conducting multivariate multilevel models, performing null models is strongly encouraged [28]. We then compared the two unconditional models using the Akaike’s Information Criterion (AIC) [29] and ANOVA tests. Lower AIC for 3-level unconditional models validated the using of 3-level models to predict stress and fatigue in our data. Finally, before performing multivariate analyses, we conducted bivariate analyses for all individual-level variables in order to determine which variables were pertinent to be included in the multivariate analysis for stress and fatigue. All variables with a p-value less or equal to 0.20 would be considered in the model. Then, we developed a 3-level multivariate model on each outcome, using AIC for variable selection. All data analyses were conducted using the R software [30].

#### Ethical approval and informed consent

The study protocol was elaborated in collaboration with the AP-HP Department of medical policy and the Department of care and of paramedical activities, and was approved after presentation to the Directorate General and the Committee on hygiene, safety and working conditions. It obtained both an agreement from the French Committee for the Protection of Persons (CPP) on 11/14/2017 and clearance from the French Data Protection Authority (CNIL) on 12/14/2017 (IDRCB N° 2017-A02939-44).

Potential participants were informed of the study through an information letter. Verbal consent was obtained by the interviewer at the beginning of each interview. Participants were guaranteed confidentiality and anonymity of responses.

## RESULTS

### Descriptive results

#### Response rate

Overall, the response rate for all included healthcare workers who answered the questionnaires at the four visits was 73.5% (510 out of 694), corresponding to 2040 responses in total for all visits. In fact, one auxiliary nurse failed to answer any of the four questions of the Karasek hierarchy questionnaire and was thus excluded from all analyses. In total, 694 healthcare workers were included at t0, 644 at t1, 578 at t2 and 556 at t3, with an overall of 2472 observations.

#### Healthcare workers characteristics **(Table 1)**

The final study sample consisted of 694 healthcare workers as follows: registered nurses (n = 384) (55.3%), auxiliary nurses (n = 299) (43.1%) and midwives (n = 11) (1.6%). Overall, the Female/Male gender ratio was 5.5, with 588 (84.7%) female respondents. The majority of healthcare workers were permanent staff members (n = 616) (88.9%) compared to temporary (n = 58) (8.4%) and contractual (n = 19) (2.7%) staff members. The average number of years of experience was 9 years (9.6), and more than a half of the respondents had supervising responsibilities (n = 365) (52.6%).

**Table 1.**
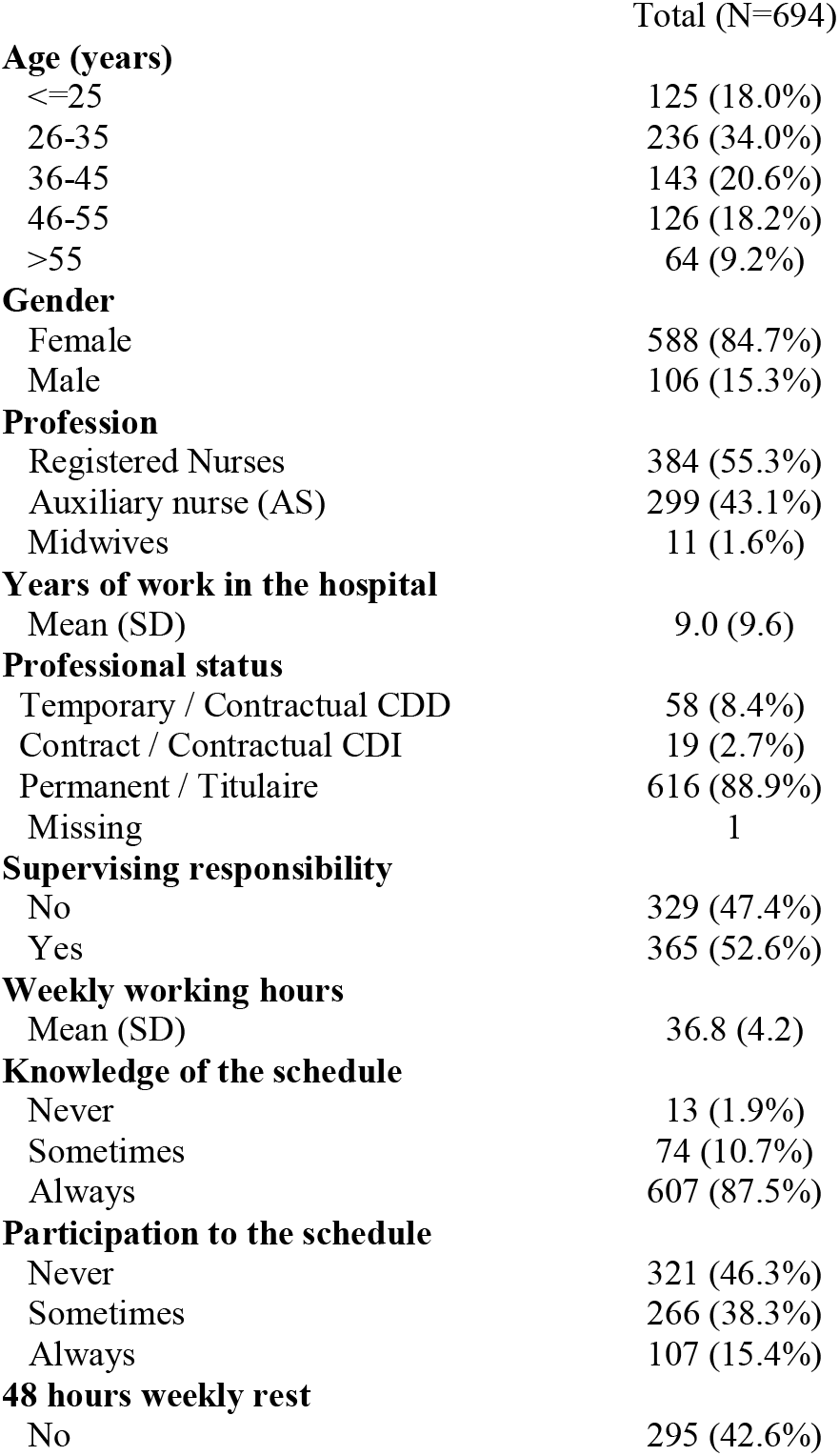
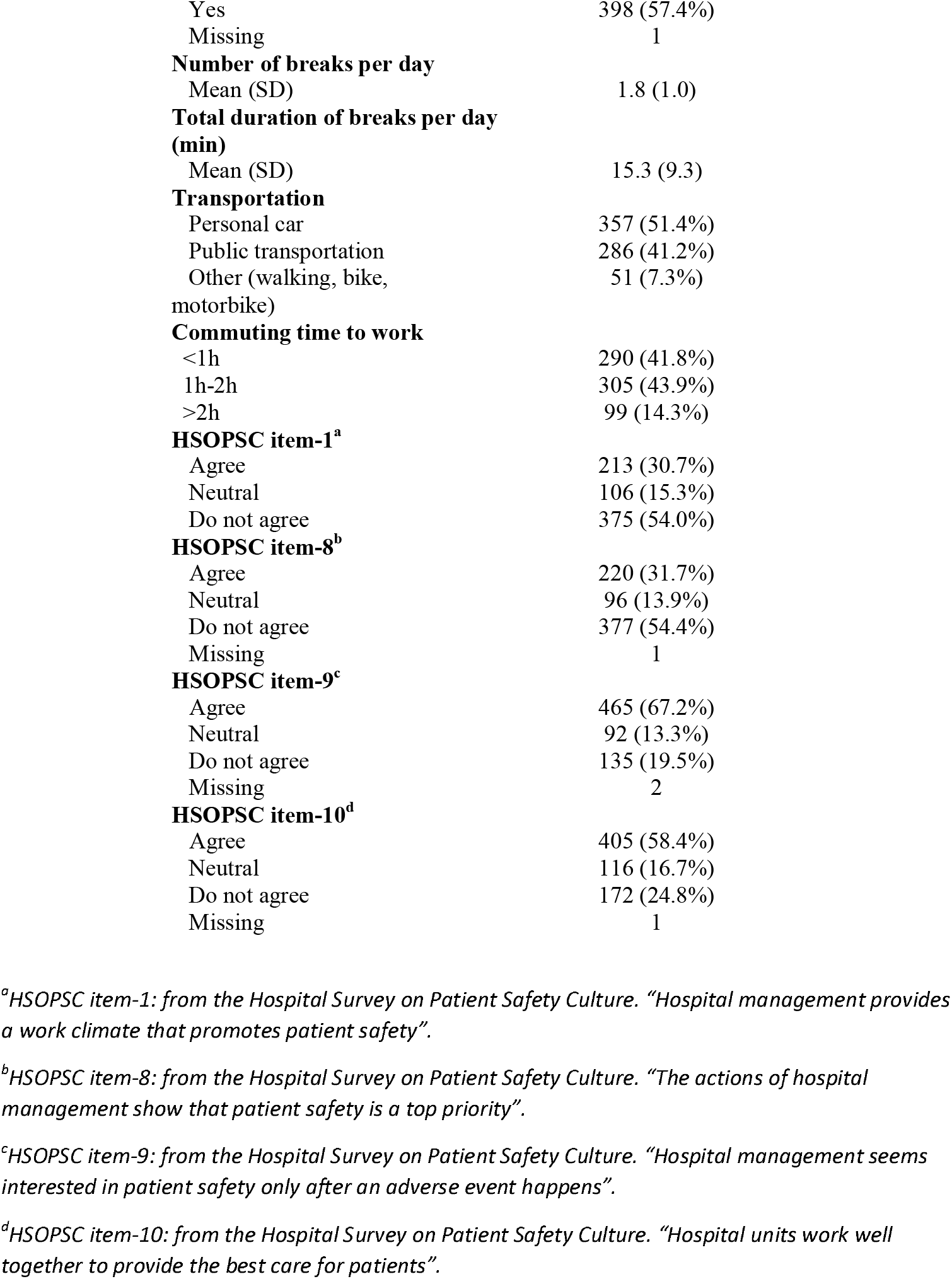
Characteristics of individuals at the time of inclusion t0.

#### Ward characteristics **(Table 2)**

In total, 32 wards were included from various medical fields as follows: 14 (43.8%) in surgery and obstetrics, 11 (34.4%) in medicine and 7 (21.9%) in intensive care units and reanimation. The average number of beds per ward was 35.5 (SD =18.5), and the proportion of double rooms was approximately 20%.

**Table 2.** Characteristics of included wards.

|  | Total (N=32) |
| --- | --- |
| **Specialty** |  |
| Surgery/Obstetrics | 14 (43.8%) |
| Medical | 11 (34.4%) |
| ICU/Reanimation | 7 (21.9%) |
| **Number of beds** |  |
| Mean (SD) | 35.5 (18.5) |
| **Proportion of double bedrooms** |  |
| Mean (SD) | 0.2 (0.2) |
| **Going outside the ward** |  |
| Never | 1 (3.2%) |
| Sometimes | 11 (35.5%) |
| Fairly Often | 14 (45.2%) |
| Always | 5 (16.1%) |
| Missing | 1 |
| **Ratio patient/physician** |  |
| Mean (SD) | 2.9 (4.0) |
| **Use of external healthcare services** |  |
| Never | 8 (25.0%) |
| Sometimes | 17 (53.1%) |
| Fairly Often | 7 (21.9%) |
| **Ratio patient/paramedics** |  |
| Mean (SD) | 0.8 (0.3) |
| **Time schedule** |  |
| 2*12h | 5 (15.6%) |
| 3*8h | 27 (84.4%) |

In participating wards, the average patient/physician ratio was 2.9 (4.0), whereas the patient/paramedics ratio was 0.8 (0.3). In the vast majority of participating wards, work was organized in 3 8-hour shifts, while 16% of wards worked in 2 12-hour shifts.

#### Mean-level change across one-year of survey

The trajectories for each participant across the total sample from baseline to the last time point are presented in supplementary material table 1 for the main variables. Significant differences among the four times of visits were observed for schedule assignment frequency, number of nightshifts on duty over the last months, irregularity of mealtimes, number of canceled breaks, number of visits to the Occupational safety and health (OSH) department, presenteeism at work while sick, and support from the hierarchy.

#### Outcome characteristics

The distribution of PSS-10 and Pichot scores in the whole sample are presented in Figure 1 respectively in a) and b). The overall mean score is equal to 16.5 (7.0) out of 40 points for stress, and 11.0 (7.9) out of 32 points for fatigue. For fatigue only, we observed an increasing trend (p = 0.028) of the means across the time of visits (supplementary table 2). For both stress and fatigue, significant differences were observed between the four hospitals (p <0.001) (supplementary table 3). Figure 2a) describes the trend of stress and fatigue level across time of visits and hospital. and significant differences were observed between the four hospitals (p <0.001) (supplementary table 3). Additional Figures on the distribution of PSS-10 and Pichot scores by hospital, are available at supplementary figures 1 and 2 respectively.

**Figure 1:**
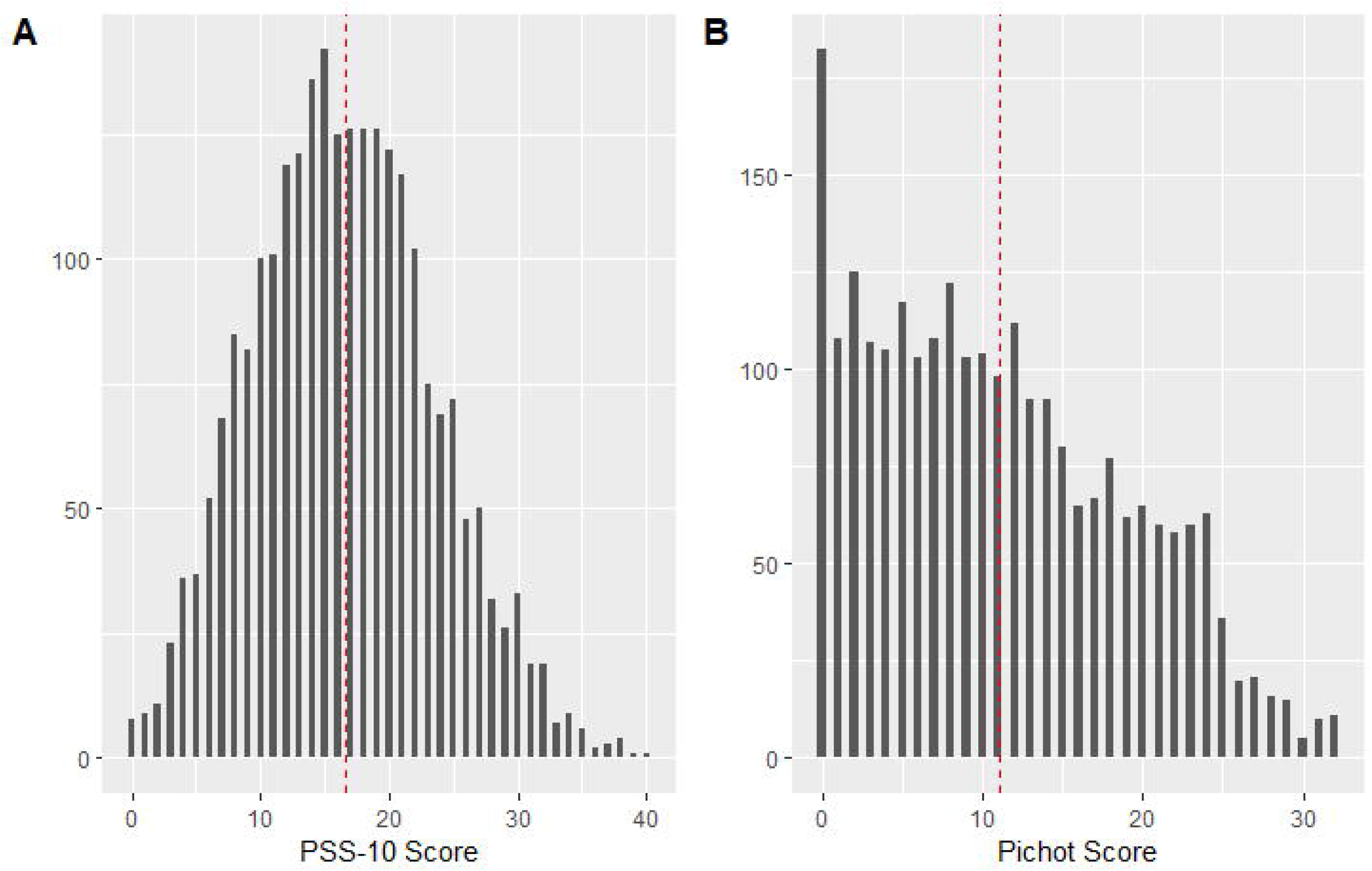
Distribution of PSS-10 and Pichot scores, respectively in a) and b) among the whole sample. The vertical dotted lines represent the mean of the PSS-10 and Pichot scores.

**Figure 2:**
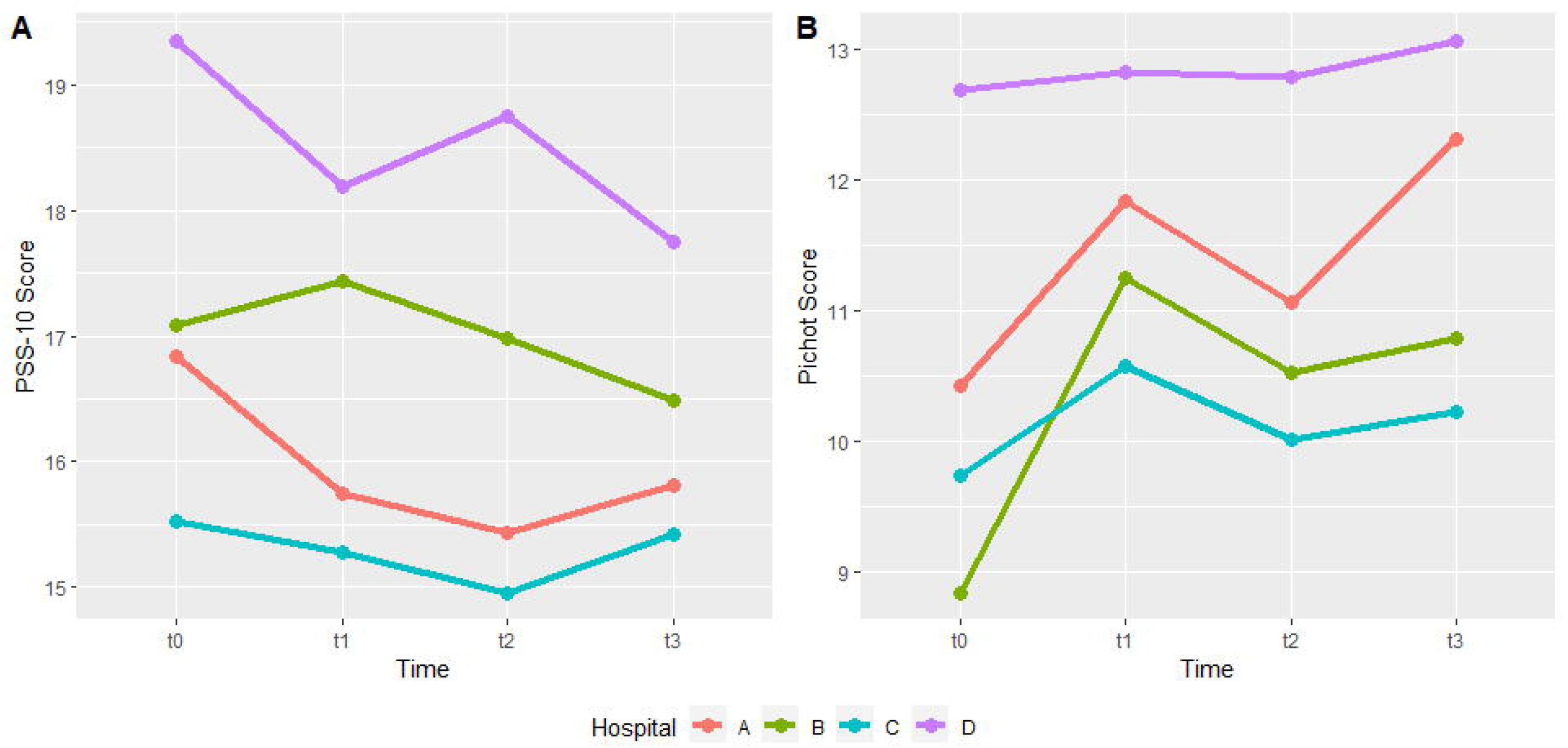
PSS-10 and Pichot scores means, respectively in a) and b), by hospital and time of visits

#### Descriptive of missing values for validated scales

A summary of missing values according to the four validated scales, by time of visits is available in supplementary table 4. Indeed, in the whole sample (n =2472), high number of missing values were observed on support from the hierarchy and perceived stress, respectively 29 and 22 values. The lowest number of missing values were observed on fatigue and work overcommitment, all two 3 missing values. We count 12 missing values in the whole sample for the support from colleague’s variable. Before proceeding with the 3-levels analyses, missing values were imputed.

### Associations of individual and organizational-level predictors with stress and fatigue

#### Unconditional models with two and three-level longitudinal modeling

Detailed results of unconditional models for both stress and fatigue, as well as their respective AICs, are shown in supplementary table 5. For both outcomes, the AICs from 3-level unconditional models are lower than those from 2-level unconditional models (15737 vs 15763 for the stress models, and 15896 vs 15922 for the fatigue models). In addition, for each outcome, we obtained significant p-values when testing 2 and 3-level models using the ANOVA test (supplementary table 5). In conclusion, the results obtained validate the using of the 3-level models to analyze both outcomes.

According to supplementary table 5, the level of stress and fatigue variance has been partitioned at all three levels. It can be easily shown that, the PSS-10 score and the Pichot score variations occurred due to temporal fluctuations (level 1, 42% for stress and 37.4% for fatigue), inter-individual heterogeneity (level 2, 52% for stress and 55.5% for fatigue) and ward-level specificities (level 3, 6% for stress and 7.2% for fatigue).

Before performing multivariate analysis, bivariate analyses of variables at healthcare workers level were conducted at inclusion (t0) for both stress and fatigue, and results are available in supplementary tables 6 and 7. All variables having p-values less or equal to 0.20 were considered and included in models before performing variable selection.

#### Risk factors associated with high levels of perceived stress

The best 3-level multivariate models identified after model selection using AIC are described in Table 3 along with model fit measures.

**Table 3:** Final 3-level models for outcomes (perceived stress and fatigue) using a stepwise approach with AIC criterion

|  | Stress (PSS-10) (n = 2422) |  |  | Fatigue (Pichot) (n = 2431) |  |  |
| --- | --- | --- | --- | --- | --- | --- |
|  | Estimates | CI | p-value | Estimates | CI | p-value |
| **Fixed effects\*** |  |  |  |  |  |  |
| Time | -0.00 | -0.16 – 0.16 | 0.957 | 0.48 | 0.31 – 0.66 | **<0.001** |
| Interviewer (ref=1)<br>2 (for hospitals B and D) | 1.62 | 0.85 – 2.40 | **<0.001** | -0.36 | -1.47 – 0.76 | 0.531 |
| **Healthcare worker-level variables** |  |  |  |  |  |  |
| **Gender (ref=Female)** |  |  |  |  |  |  |
| Male | -0.83 | -1.73 – 0.06 | 0.068 | -1.52 | -2.63 – -0.40 | **0.008** |
| **Age (ref=<25)** |  |  |  |  |  |  |
| 26-35 |  |  |  | 1.26 | 0.09 – 2.44 | **0.035** |
| 36-45 |  |  |  | 0.50 | -0.79 – 1.78 | 0.450 |
| 46-55 |  |  |  | -1.01 | -2.37 – 0.35 | 0.147 |
| >55 |  |  |  | -2.83 | -4.49 – -1.17 | **0.001** |
| **Commuting time to work (ref=>1h)** |  |  |  |  |  |  |
| >2h |  |  |  | 1.82 | 0.80 – 2.83 | **<0.001** |
| 1h-2h |  |  |  | 0.30 | -0.37 – 0.96 | 0.382 |
| **Personal life event (ref=No)** |  |  |  |  |  |  |
| Yes, negative | 2.27 | 1.81 – 2.74 | **<0.001** |  |  |  |
| Yes, positive | 0.46 | -0.19 – 1.12 | 0.164 |  |  |  |
| **Professional life event (ref=No)** |  |  |  |  |  |  |
| Yes, negative | 1.62 | 1.11 – 2.12 | **<0.001** |  |  |  |
| Yes, positive | 0.40 | -0.34 – 1.14 | 0.289 |  |  |  |
| **Break canceled (ref=Never)** |  |  |  |  |  |  |
| Almost never | 1.04 | 0.27 – 1.82 | **0.008** |  |  |  |
| Quite often | 1.54 | 0.77 – 2.32 | **<0.001** |  |  |  |
| Very often | 1.78 | 0.92 – 2.65 | **<0.001** |  |  |  |
| **Professional status (ref=Temporary CDD)** |  |  |  |  |  |  |
| Contract (Contractuel CDI) |  |  |  | 2.23 | -0.53 – 4.99 | 0.114 |
| Permanent (Titulaire) |  |  |  | 0.45 | -1.04 – 1.93 | 0.554 |
| **Knowing work schedule in advance (ref= Never)** |  |  |  |  |  |  |
| Sometimes | -2.30 | -4.81 – 0.22 | 0.074 |  |  |  |
| Always | -1.87 | -4.23 – 0.49 | 0.121 |  |  |  |
| **Support from colleagues – Karasek score** | -0.12 | -0.24 – -0.01 | **0.035** |  |  |  |
| **Support from hierarchy – Karasek score** | -0.23 | -0.33 – -0.13 | **<0.001** | -0.34 | -0.45 – -0.24 | **<0.001** |
| **HSOPSC item-1<sup>a</sup> (ref=Agree)** |  |  |  |  |  |  |
| Neutral | 0.00 | -1.01 – 1.02 | 0.995 | 0.70 | -0.52 – 1.91 | 0.260 |
| Not Agree | 0.81 | -0.10 – 1.71 | 0.082 | 1.76 | 0.81 – 2.70 | **<0.001** |
| **HSOPSC item-8<sup>b</sup> (ref=Agree)** |  |  |  |  |  |  |
| Neutral | 0.46 | -0.58 – 1.51 | 0.385 |  |  |  |
| Not Agree | 0.96 | 0.07 – 1.84 | **0.034** |  |  |  |
| **Work overcommitment – Siegrist score** | 0.71 | 0.63 – 0.78 | **<0.001** | 0.65 | 0.56 – 0.73 | **<0.001** |
| **Presence at work while sick (ref=Never)** |  |  |  |  |  |  |
| Almost never | 0.04 | -0.48 – 0.57 | 0.871 | 0.63 | 0.04 – 1.22 | **0.037** |
| Quite often | 1.04 | 0.45 – 1.63 | **0.001** | 2.48 | 1.82 – 3.15 | **<0.001** |
| Very often | 1.61 | 0.55 – 2.66 | **0.003** | 4.03 | 2.84 – 5.22 | **<0.001** |
| **Ward-level variables** |  |  |  |  |  |  |
| **Medical specialty (ref=surgery/obstetrics)** |  |  |  |  |  |  |
| Medicine | -0.36 | -1.30 – 0.59 | 0.459 | 1.59 | 0.24 – 2.94 | **0.021** |
| ICU/Reanimation | -1.08 | -2.01 – -0.15 | **0.023** | -0.24 | -1.60 – 1.13 | 0.733 |
| **Number of beds per ward** | -0.03 | -0.05 – -0.01 | **0.009** | -0.04 | -0.07 – -0.00 | **0.024** |
| **Going outside the ward (ref = Never)** |  |  |  |  |  |  |
| Sometimes | 1.28 | -0.87 – 3.43 | 0.243 | 0.98 | -2.09 – 4.04 | 0.533 |
| Often | 1.53 | -0.70 – 3.76 | 0.178 | 0.86 | -2.30 – 4.03 | 0.592 |
| Always | 1.87 | -0.44 – 4.18 | 0.112 | 2.59 | -0.70 – 5.89 | 0.123 |
| **Use of interim nurses (ref = Never)** |  |  |  |  |  |  |
| Sometimes | 0.05 | -0.81 – 0.91 | 0.915 | -0.36 | -1.59 – 0.88 | 0.571 |
| Often | 0.96 | -0.14 – 2.06 | 0.088 | 1.96 | 0.35 – 3.56 | **0.017** |
| **Random Effects $\sigma^2(\sigma)^*$** |  |  |  |  |  |  |
| Level 1 – Time |  | 17.80 (4.23) |  |  | 22.04 (4.69) |  |
| Level 2 – Healthcare worker |  | 12.30 (3.51) |  |  | 19.69 (4.44) |  |
| Level 3 – Ward |  | 0.00 (0.00) |  |  | 0.50 (0.70) |  |
| <sup>d</sup>Marginal R<sup>2</sup> / Conditional R<sup>2</sup> |  | 0.507 / NA |  |  | 0.299 / 0.634 |  |
| AIC |  | 14743.16 |  |  | 15433.57 |  |

- <sup>a</sup> HSOPSC item-1: item-1 from the Hospital Survey on Patient Safety Culture. “Hospital management provides a work climate that promotes patient safety”.
- <sup>b</sup> *HSOPSC item-8: item-8 from the Hospital Survey on Patient Safety Culture. “The actions of hospital management show that patient safety is a top priority”.* - <sup>c</sup> ICC intra-class correlation coefficient, AIC Akaike Information Criterion - <sup>d</sup> The marginal R<sup>2</sup> considers only the variance of the fixed effects, while the conditional R<sup>2</sup> takes both the fixed and random effects into account - \* In multilevel models, fixed effects are usually equivalent to the regression coefficients, while random effects usually account for the underlying structure of the data and characterized by estimates of variability ( $\sigma^2(\sigma)$ ). Fixed effects can be interpreted as slopes in the traditional sense - -

High perceived stress was best explained by:

- Individual level variables: negative life events, breaks frequently canceled, lack of support from hierarchy and colleagues, low perceived safety culture in the hierarchy, work overcommitment, presenteeism at work while sick;
- Ward-level variables: medical specialty (with less stress in intensive care units), and number of beds (with less stress in larger wards).

Risk factors associated with high levels of fatigue:

High fatigue level was best explained by:

- Time-level variable: time of visit, with an increasing trend
- Individual level variables: gender (with less fatigue in males), long commuting time to work, lack of hierarchical support, low perceived safety culture in the hierarchy, work overcommitment, presenteeism while sick at work.
- Ward-level variables: medical specialty (with more fatigue in medical wards), high rates of interim use and number of beds in the ward (with less fatigue in larger wards).

## DISCUSSION

### Main findings

In this longitudinal study, we underlined the role of several individual and organizational factors in the stress and fatigue of healthcare workers. One purpose of longitudinal three-level modeling is to assess environmental wards (Level 3) influences on healthcare workers average change (Level 2) over time (Level 1). In particular, a hierarchy providing low support and with low safety culture, work overcommitment, and presenteeism while sick, and working in smaller wards were identified as predictors of both high stress and fatigue. In addition, high frequency of break cancellation, negative life event (whether personal or professional) and low support from the colleagues are specific predictors of high stress. However, long commuting duration, frequent use of interim staff and working in a medical ward were associated with high level of fatigue.

In our data, the proportion of healthcare workers with extreme fatigue and extreme perceived stress are very high, respectively 37% and 30.5%. These proportions were measured considering the cut-off of 22 for extreme fatigue [31] and 27 for extreme perceived stress [32]. We using the PSS-10 for measured extreme perceived stress and although it is not a diagnostic instrument and there is no predetermined cut-off for PSS score [24], some studies used PSS scores of 0–13, 14–26, and 27–40 points to assess low, moderate, and high perceived stress, respectively [32]. Extreme fatigue is experienced mostly among healthcare workers aged between 26 and 45 years old (32.6%), females (93%), and working in surgery/obstetrics wards. Even though the multivariate model shows high fatigue in medical ward, healthcare workers in surgery/obstetrics wards experienced individually an extreme fatigue due to the amount of work. Regarding extreme stress, the most at-risk groups were also healthcare workers between 26 and 45 years old (this group alone account for 58.5%), female (89.6%) and working in surgery/obstetrics wards. The multi-level model of stress confirms this result by showing lower stress for healthcare workers working in intensive care unit/reanimation wards than in surgery/obstetrics wards. For both extreme fatigue and extreme stress, there is no a significant trend over time.

### Comparison with the literature

Many of our findings are consistent with those reported in previous studies investigating the determinants of stress or fatigue in healthcare workers.

In particular, the influence of the lack of hierarchical support on both stress and fatigue was underlined in an earlier French study [15]. This same study also showed higher fatigue in small to medium hospital wards, and in work environments where staff frequently had to go outside the ward, as well as lower energy levels and more frequent sleep difficulties when use of interim staff was frequent, consistently with our results [15]. Safety climate perceptions were found to be significantly related to healthcare worker stress and in several recent studies [33,34]. Our finding that work overcommitment, as measured by the ERI questionnaire, and presenteeism while sick, another indicator of overcommitment, were significant predictors of high stress and fatigue in healthcare workers is supported by a recent French study in which overcommitment was found to favor emotional exhaustion and increase the risk of burnout in French healthcare workers [35].

Other factors previously reported in the literature as associated with stress, but not in our results, were low social support [13,36], rearing children, work on rotation, chronic medical illnesses [12] and being unsatisfied at work [1]. Regarding fatigue, another associated factor previously reported in the literature but not in our results was work over longer shifts (12h versus 8h) [15,37]. This could be explained by a potential lack of power due to the sample size as in our sample, there were very few wards with 12-hours shift compared to those with an 8-hours shift.

### Strengths

First, one strength of this study is its longitudinal nature. A few studies explored stress and fatigue longitudinally [17,38], however the majority of currently available studies are cross-sectional. Furthermore, the high response rate of wards and healthcare workers, as well as the large sample size and the inclusion of wards of different size and activity, represent a strength of this study. In addition, the large panel of socio-demographics, health and occupational characteristics of healthcare workers collected over time allow to perform robust and well-adjusted multivariate analysis.

Second, stress and fatigue were explored together. To our knowledge, no previous study had proposed a single model to identify factors associated with high levels of combined stress and fatigue while accounting for time in hospital setting. In fact, a previous study conducted in French ICU attempted to predict stress and fatigue using demographic and occupational was based on cross-sectional survey [15].

Finally, the power of the model used, which takes into account the complexity of the data, namely the longitudinal design and the multi wards collection of the data. In recent years, these types of models have been frequently used [39,40] considering the idea that longitudinal data could be analyzed at three levels of nesting (e.g., repeated measures [Level 1], collected across individuals [Level 2], and within different wards [Level 3]).

## Limitations

However, our study has some limitations. First, only 4 hospitals in Paris-area were included, which is not representative for other areas of France. In fact, only services in public Paris-area hospitals were included in the study, so results may not be generalizable to wards in private hospitals or outside the Paris region, Future studies including healthcare workers and more hospitals from other cities in France are needed to verify the results generated in this study.

Second, due to the in-person interview, a risk of bias could be present due to the due to discomfort from having to reply face-to-face to some sensitive questions; however, ensuring anonymity of the participants was used to minimize such bias. Another possible source of bias in data collection is the presence of two different interviewers assigned to two hospitals each. However, we were able to consider this bias as we included this interviewer-related variable into the multi-level model. In the fatigue model, there was a significant interviewer effect, with higher stress in hospital B and D. However, interviewer effect was not significant in the stress model.

Third, we were not able to investigate stress and fatigue outcomes for the physicians given low response rates, as questions regarding work organization were less adequate than for nurses. Therefore, physicians were excluded from our sample of healthcare workers.

### Insights for designing potential interventions

From these models, we can identify some areas for improvement to better prevent stress and fatigue of healthcare worker: (1) perception of the hierarchy (lack of support from the hierarchy, low perceived safety culture of the hierarchy), (2) work overcommitment and (3) presenteeism at work while sick). Cancellation of breaks and support from colleagues were also found as significant as specific predictors for stress level. For fatigue specifically, long commuting duration and use of external staff are also identified as predictors. Mutual and specifics preventive programs for reducing stress and fatigue of healthcare workers could be implemented in order to reduce this burden, targeting on the most at-risk groups. Other variables (medical specialty, number of beds) are inherent to one given ward; hence they have less utility for interventions to reduce perceived stress and fatigue.

## CONCLUSION

This research question is important given the influence on quality of patient care of high stress work environments [7]. Our results may (1) help identify at-risk healthcare workers and wards, where interventions to reduce stress and fatigue could be focused. (2) These interventions could include manager training to favor better staff support and overall safety culture among healthcare workers.

## Data Availability

All data produced in the present study are available upon reasonable request to the authors

## Contributors

Oumou Salama Daouda: Methodology, Software, Formal analysis, Writing -Original Draft, Writing - Review & Editing, Visualization, Validation

René Sosata Bun: Methodology, Formal analysis, Writing - Original Draft, Writing - Review & Editing, Visualization, Validation

Karim Ait Bouziad: Methodology, Formal analysis, Writing - Review & Editing, Visualization, Validation

Katiuska Miliani: Conceptualization, Writing - Review & Editing, Visualization, Validation Anastasia Essa-Eworo: Investigation

Florence Espinasse: Conceptualization, Supervision, Project administration

Delphine Seytre: Conceptualization, Supervision, Project administration

Anne Casseta: Conceptualization, Supervision, Project administration

Simone Nérome: Conceptualization, Supervision, Project administration

Adelaide Nascimento: Conceptualization, Visualization, Funding acquisition, Validation

Pascal Astagneau: Conceptualization, Methodology, Formal analysis, Writing - Review & Editing, Visualization, Supervision, Funding acquisition, Validation

Laura Temime: Conceptualization, Methodology, Formal analysis, Writing - Review & Editing, Visualization, Supervision, Funding acquisition, Validation

Mounia N Hocine: Conceptualization, Methodology, Formal analysis, Writing - Review & Editing, Visualization, Supervision, Funding acquisition, Validation

## Acknowledgments

The sponsor was Assistance Publique – Hôpitaux de Paris (Délégation à la Recherche Clinique et à l’Innovation). The authors would like to thank Zahoua Tina Habbal, the AP-HP and all participating wards and healthcare workers for their time and their contribution to this study.

## Conflicts of interest

The authors have no conflicts of interest to declare that are relevant to the content of this article.

## Funding sources

The study was publicly funded through the Research on the Evaluation of the Healthcare System Performance Program, Ministry of Health, France (grant No. PREPS- 16-261)

## Notes

### Competing Interest Statement

The authors have declared no competing interest.

### Author Declarations

The study protocol was elaborated in collaboration with the AP-HP Department of medical policy and the Department of care and of paramedical activities, and was approved after presentation to the Directorate General and the Committee on hygiene, safety and working conditions. It obtained both an agreement from the French Committee for the Protection of Persons (CPP) on 11/14/2017 and clearance from the French Data Protection Authority (CNIL) on 12/14/2017 (IDRCB Number 2017-A02939-44).

