## Supplemental file for "A multilevel approach to individual and organizational predictors of stress and fatigue among healthcare workers of a university hospital: A longitudinal study"

**Table of contents**

**Supplementary figure 1:** Distribution of PSS-10 score. by hospital. The vertical dashed lines represent the means of the PSS-10 scores by hospital

**Supplementary figure 2:** Distribution of Pichot score by hospital. The vertical dashed lines represent the means of the Pichot scores by hospital

**Supplementary table 1**. Characteristics of individuals and missing values at the times of visits (t0, t1, t2 and t3)

**Supplementary table 2**:  Means and ranges of outcomes variables (PSS-10 and Pichot scores), by time of visits

**Supplementary table 3**: Outcomes (PSS-10 and Pichot scores), and missing values by hospital

**Supplementary table 4:** Summary of missing values according to validated scales, by time of visits

**Supplementary table 5**: Unconditional 2 and 3-level models for outcomes of perceived stress and fatigue

**Supplementary table 6**: Bivariate analysis (linear regressions) of variables at healthcare workers level for stress level (PSS-10 score) at inclusion. Variables were sorted according to their p-values. Variables in bold (p_value > 0 .20) were not considered in the multivariate analysis

**Supplementary table 7**: Bivariate analysis (linear regressions) of variables at healthcare workers level for fatigue level (Pichot score) at inclusion. Variables were sorted according to their p-values. Variables in bold (p_value > 0 .20) were not considered in the multivariate analysis, expected the "commuting time to work" variable.

**Supplementary figure 1:** Distribution of PSS-10 score. by hospital. The vertical dashed lines represent the means of the PSS-10 scores by hospital

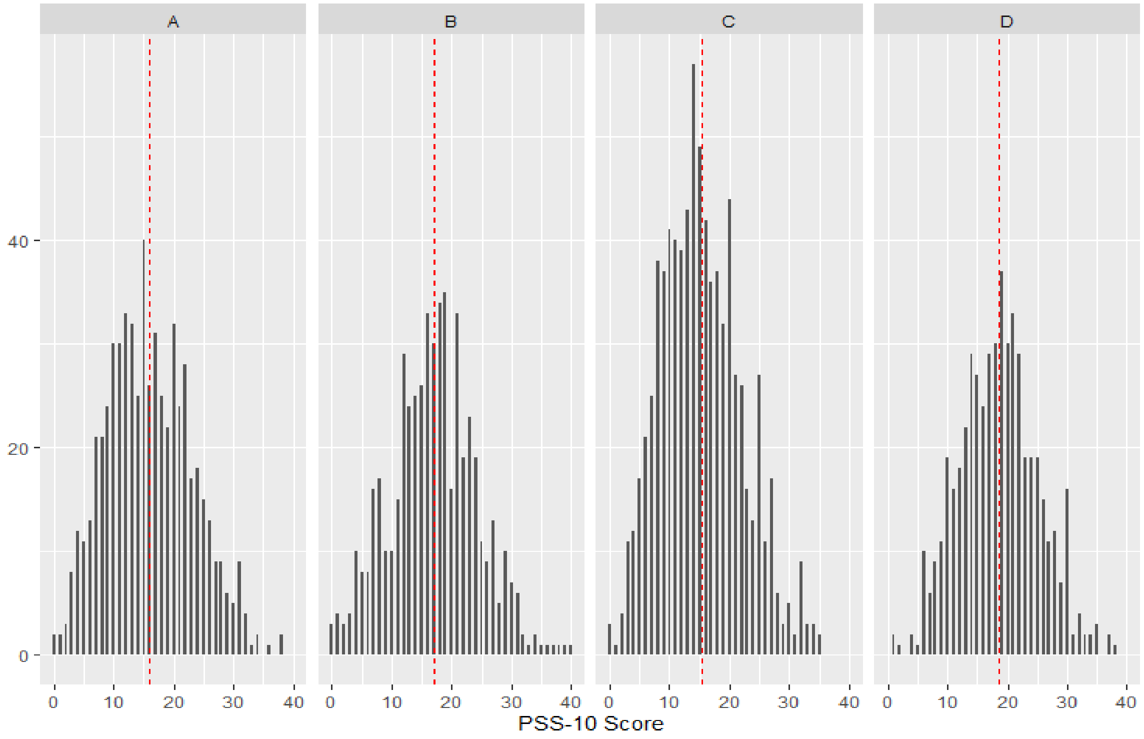


**Supplementary figure 2:** Distribution of Pichot score by hospital. The vertical dashed lines represent the means of the Pichot scores by hospital

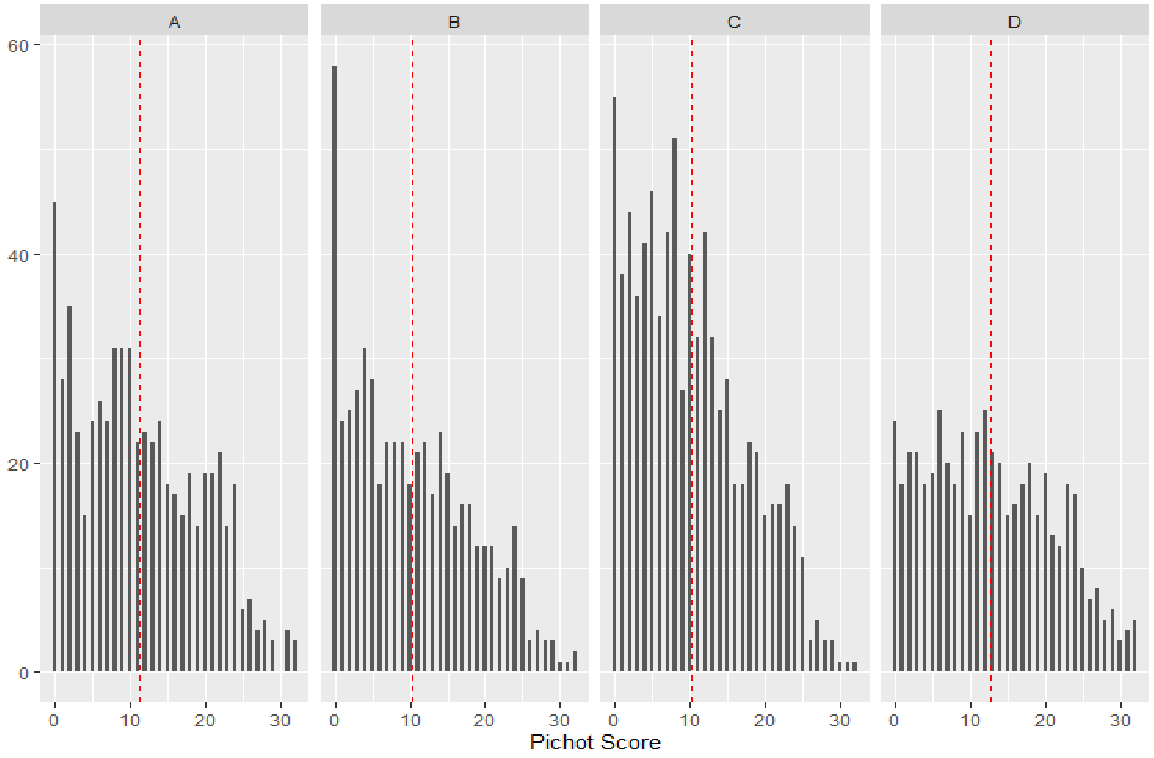

**Supplementary table 1.** Characteristics of individuals and missing values at the times of visits (t0, t1, t2 and t3)

Two-sided ANOVA tests were performed for continuous variables and Chi square tests were performed for qualitative variables

| | t0 (N=694) | t1 (N=644) | t2 (N=578) | t3 (N=556) | Total (N=2472) | p value |
|---|---|---|---|---|---|---|
| **Work schedule of last months** | | | | | | 0.636 |
| Daily | 458 (66.0%) | 436 (67.7%) | 381 (65.9%) | 357 (64.2%) | 1632 (66.0%) | |
| Nightly | 198 (28.5%) | 185 (28.7%) | 170 (29.4%) | 168 (30.2%) | 721 (29.2%) | |
| Day and Night | 38 (5.5%) | 23 (3.6%) | 27 (4.7%) | 31 (5.6%) | 119 (4.8%) | |
| **Schedule assignment frequency** | | | | | | < 0.001 |
| Mostly | 76 (11.0%) | 47 (7.3%) | 23 (4.0%) | 36 (6.5%) | 182 (7.4%) | |
| Always | 618 (89.0%) | 597 (92.7%) | 555 (96.0%) | 519 (93.5%) | 2289 (92.6%) | |
| Missing | 0 | 0 | 0 | 1 | 1 | |
| **Nightshift/duty on last months** | | | | | | < 0.001 |
| No | 564 (81.4%) | 380 (59.1%) | 324 (56.2%) | 319 (57.7%) | 1587 (64.4%) | |
| Yes | 129 (18.6%) | 263 (40.9%) | 252 (43.8%) | 234 (42.3%) | 878 (35.6%) | |
| Missing | 1 | 1 | 2 | 3 | 7 | |
| **Number of nightshift/duties** | | | | | | < 0.001 |
| Mean (SD) | 0.6 (1.7) | 1.3 (2.2) | 1.5 (2.3) | 1.5 (2.4) | 1.2 (2.2) | |
| Missing | 4 | 5 | 2 | 3 | 14 | |
| **Work schedule variation** | | | | | | 0.762 |
| Never | 404 (58.2%) | 360 (55.9%) | 334 (57.8%) | 319 (57.4%) | 1417 (57.3%) | |
| Fairly often | 122 (17.6%) | 123 (19.1%) | 94 (16.3%) | 100 (18.0%) | 439 (17.8%) | |
| Almost Never | 146 (21.0%) | 132 (20.5%) | 133 (23.0%) | 114 (20.5%) | 525 (21.2%) | |
| Very often | 22 (3.2%) | 29 (4.5%) | 17 (2.9%) | 23 (4.1%) | 91 (3.7%) | |
| **Overtime hours** | | | | | | 0.098 |
| Never | 238 (34.3%) | 209 (32.5%) | 177 (30.6%) | 179 (32.2%) | 803 (32.5%) | |
| Fairly often | 205 (29.5%) | 209 (32.5%) | 208 (36.0%) | 189 (34.0%) | 811 (32.8%) | |
| Almost Never | 173 (24.9%) | 180 (28.0%) | 149 (25.8%) | 139 (25.0%) | 641 (25.9%) | |

| | | | | | | |
|---|---|---|---|---|---|---|
| Very often | 78 (11.2%) | 46 (7.1%) | 44 (7.6%) | 49 (8.8%) | 217 (8.8%) | |
| **Irregularity of meal time** | | | | | | 0.047 |
| Never | 41 (5.9%) | 51 (7.9%) | 34 (5.9%) | 37 (6.7%) | 163 (6.6%) | |
| Fairly often | 191 (27.6%) | 182 (28.3%) | 188 (32.5%) | 175 (31.5%) | 736 (29.8%) | |
| Almost Never | 62 (8.9%) | 55 (8.5%) | 63 (10.9%) | 69 (12.4%) | 249 (10.1%) | |
| Very often | 399 (57.6%) | 356 (55.3%) | 293 (50.7%) | 275 (49.5%) | 1323 (53.5%) | |
| Missing | 1 | 0 | 0 | 0 | 1 | |
| **Number of canceled breaks** | | | | | | < 0.001 |
| Never | 48 (6.9%) | 75 (11.6%) | 67 (11.6%) | 65 (11.7%) | 255 (10.3%) | |
| Fairly often | 279 (40.2%) | 266 (41.3%) | 229 (39.6%) | 221 (39.7%) | 995 (40.3%) | |
| Almost Never | 129 (18.6%) | 128 (19.9%) | 152 (26.3%) | 162 (29.1%) | 571 (23.1%) | |
| Very often | 238 (34.3%) | 175 (27.2%) | 130 (22.5%) | 108 (19.4%) | 651 (26.3%) | |
| **Number of visits to the Occupational safety and health (OSH) department** | | | | | | < 0.001 |
| Mean (SD) | 0.3 (0.5) | 0.2 (0.5) | 0.1 (0.4) | 0.2 (0.5) | 0.2 (0.5) | |
| **Personal life events** | | | | | | 0.148 |
| No | 408 (58.8%) | 383 (59.6%) | 350 (60.6%) | 337 (60.6%) | 1478 (59.8%) | |
| Yes, negative | 217 (31.3%) | 192 (29.9%) | 166 (28.7%) | 141 (25.4%) | 716 (29.0%) | |
| Yes, positive | 69 (9.9%) | 68 (10.6%) | 62 (10.7%) | 78 (14.0%) | 277 (11.2%) | |
| Missing | 0 | 1 | 0 | 0 | 1 | |
| **Professional life events** | | | | | | 0.557 |
| No | 488 (70.5%) | 463 (71.9%) | 390 (67.6%) | 381 (68.6%) | 1722 (69.8%) | |
| Yes, negative | 151 (21.8%) | 139 (21.6%) | 137 (23.7%) | 123 (22.2%) | 550 (22.3%) | |
| Yes, positive | 53 (7.7%) | 42 (6.5%) | 50 (8.7%) | 51 (9.2%) | 196 (7.9%) | |
| Missing | 2 | 0 | 1 | 1 | 4 | |
| **Presence at work while sick** | | | | | | 0.005 |
| Never | 159 (22.9%) | 192 (29.9%) | 122 (21.1%) | 154 (27.7%) | 627 (25.4%) | |
| Fairly often | 218 (31.4%) | 178 (27.7%) | 174 (30.2%) | 148 (26.7%) | 718 (29.1%) | |
| Almost Never | 275 (39.6%) | 242 (37.7%) | 251 (43.5%) | 235 (42.3%) | 1003 (40.6%) | |
| Very often | 42 (6.1%) | 30 (4.7%) | 30 (5.2%) | 18 (3.2%) | 120 (4.9%) | |
| Missing | 0 | 2 | 1 | 1 | 4 | |
| **Marital status** | | | | | | 0.371 |

| | | | | | | |
|---|---|---|---|---|---|---|
| Couple | 382 (55.0%) | 360 (55.9%) | 336 (58.3%) | 328 (59.4%) | 1406 (57.0%) | |
| Single | 312 (45.0%) | 284 (44.1%) | 240 (41.7%) | 224 (40.6%) | 1060 (43.0%) | |
| Missing | 0 | 0 | 2 | 4 | 6 | |
| **Commuting time to work** | | | | | | 0.109 |
| <1 h | 290 (41.8%) | 270 (41.9%) | 253 (43.8%) | 243 (43.7%) | 1056 (42.7%) | |
| >2h | 99 (14.3%) | 91 (14.1%) | 54 (9.3%) | 60 (10.8%) | 304 (12.3%) | |
| 1-2 h | 305 (43.9%) | 283 (43.9%) | 271 (46.9%) | 253 (45.5%) | 1112 (45.0%) | |
| **Support from colleagues (Karasek score)** | | | | | | 0.476 |
| Mean (SD) | 13.2 (2.0) | 13.1 (2.0) | 13.1 (2.0) | 13.0 (2.0) | 13.1 (2.0) | |
| **Support from hierarchy (Karasek score)** | | | | | | < 0.001 |
| Mean (SD) | 11.7 (2.7) | 11.5 (2.5) | 11.4 (2.6) | 11.1 (2.8) | 11.4 (2.7) | |
| **Work overcommitment (Siegrist score)** | | | | | | 0.558 |
| Mean (SD) | 15.5 (2.7) | 15.4 (2.7) | 15.3 (2.6) | 15.3 (2.6) | 15.4 (2.7) | |

**Supplementary table 2:** Means and ranges of outcomes variables (PSS-10 and Pichot scores), by time of visits

| | t0 (N=694) | t1 (N=644) | t2 (N=578) | t3 (N=556) | Total (N=2472) | p_value |
|---|---|---|---|---|---|---|
| **PSS-10 score** | | | | | | 0.126 |
| Mean (SD) | 17.0 (7.0) | 16.5 (7.0) | 16.3 (7.0) | 16.2 (7.1) | 16.5 (7.0) | |
| Range | 0.0 - 38.0 | 0.0 - 38.0 | 1.0 - 40.0 | 0.0 - 39.0 | 0.0 - 40.0 | |
| **Pichot score** | | | | | | 0.028 |
| Mean (SD) | 10.4 (7.8) | 11.5 (8.0) | 10.9 (7.9) | 11.4 (7.9) | 11.0 (7.9) | |
| Range | 0.0 - 32.0 | 0.0 - 32.0 | 0.0 - 32.0 | 0.0 - 32.0 | 0.0 - 32.0 | |

**Supplementary table 3:** Outcomes (PSS-10 and Pichot scores), and missing values by hospital

| | A (N=610) | B (N=538) | C (N=801) | D (N=523) | Total (N=2472) | p_value |
|---|---|---|---|---|---|---|
| **PSS-10 score** | | | | | | < 0.001 |
| Mean (SD) | 16.0 (7.2) | 17.0 (7.2) | 15.3 (6.8) | 18.6 (6.6) | 16.5 (7.0) | |
| Missing | 4 | 12 | 2 | 4 | 22 | |
| **Pichot score** | | | | | | < 0.001 |
| Mean (SD) | 11.4 (8.0) | 10.3 (7.9) | 10.1 (7.3) | 12.8 (8.3) | 11.0 (7.9) | |
| Missing | 0 | 0 | 2 | 1 | 3 | |

**Supplementary table 4:** Summary of missing values according to validated scales, by time of visits

| | **t0** | **t1** | **t2** | **t3** | **Total** |
|---|---|---|---|---|---|
| PSS-10 score - Stress | 7 | 3 | 5 | 7 | 22 |
| Pichot score - Fatigue | 1 | 0 | 2 | 0 | 3 |
| Karasek score - Support from colleagues | 6 | 4 | 1 | 1 | 12 |
| Karasek score - Support from hierarchy | 23 | 5 | 1 | 0 | 29 |
| Siegrist score - Work overcommitment | 1 | 1 | 0 | 1 | 3 |

**Supplementary table 5:** Unconditional 2 and 3-level models for outcomes of perceived stress and fatigue

| | Stress – PSS-10 Score | | Fatigue – Pichot Score | |
|---|---|---|---|---|
| | **2-level** | **3-level** | **2-level** | **3-level** |
| Intercept | 16.7(0.23) | 16.9 (0.38) | 11.2 (0.25) | 11.4 (0.5) |
| **Random effects - $\sigma^2(\sigma)$*** | | | | |
| Level 1 – Time | 28.9 (5.3) | 20.9 (4.6) | 23.9 (4.8) | 23.8 (4.9) |
| Level 2 – Healthcare worker | 28.94 (5.3) | 25.9 (5.1) | 39.12 (6.3) | 35.1 (6) |
| Level 3 – Ward | | 3.2 (1.8) | | 4.6 (2.1) |
| ICC[a] Ward | | | | |
| ICC[a] Healthcare worker within ward | | | | |
| AIC[a] | 15762.86 | 15737.24 | 15922.34 | 15896.45 |
| ANOVA test p-value | < 2e-16 | | 2.25 x 10e-07 | |

[a] ICC intra-class correlation coefficient, AIC Akaike Information Criterion

**Supplementary table 6:** Bivariate analysis (linear regression) of variables at healthcare workers level on stress level (PSS-10 score) at inclusion. Variables were sorted according to their p-values. Variables in bold (p_value > 0 .20) were not considered in the multivariate analysis

| **Variables** | **p_value** |
|---|---|
| Work overcommitment (Siegrist score) | 7.01 e-46 |
| Professional life events | 1.86 e-16 |
| HSOPSC item-1[a] | 3.36 e-16 |
| HSOPSC item-8[b] | 3.20 e-11 |
| Support from hierarchy (Karasek score) | 2.10 e-10 |
| Presence at work while sick | 2.76 e-10 |
| Personal life events | 8.44 e-09 |
| Breaks cancelled | 1.44 e-05 |
| HSOPSC item-10[d] | 1.88 e-05 |
| 48 hours weekly rest | 2.19 e-05 |
| Knowledge of the schedule | 7.42 e-05 |
| Gender | 0.0005 |
| Support from hierarchy (Karasek score) | 0.0006 |
| Number of breaks per day | 0.0038 |
| Work schedule variation | 0.0070 |
| HSOPSC item-9[c] | 0.0080 |
| Age | 0.0140 |
| Transportation | 0.0325 |
| Total duration of breaks per day (min) | 0.0588 |
| Number of nightshift/duties | 0.0680 |
| Number of children (between 0 and 10 years old) | 0.0754 |
| Irregularity of meal time | 0.1035 |
| Years of work in the hospital | 0.1062 |

| | |
|---|---|
| Overtime hours | 0.1077 |
| Marital status | 0.1555 |
| Supervising responsibility | 0.1650 |
| **Participation to the schedule** | 0.2116 |
| **Work schedule of last months** | 0.2815 |
| **Number of visits to the Occupational safety and health (OSH) department** | 0.4099 |
| **Commuting time to work** | 0.5311 |
| **Nightshift/duty on last months** | 0.6609 |
| **Weekly working hours** | 0.7558 |
| **Profession** | 0.7606 |
| **Schedule assignment frequency** | 0.8019 |
| **Professional status** | 0.8250 |

[a]*HSOPSC item-1: from the Hospital Survey on Patient Safety Culture. "Hospital management provides a work climate that promotes patient safety".*

[b]*HSOPSC item-8: from the Hospital Survey on Patient Safety Culture. "The actions of hospital management show that patient safety is a top priority".*

[c]*HSOPSC item-9: from the Hospital Survey on Patient Safety Culture. "Hospital management seems interested in patient safety only after an adverse event happens".*

[d]*HSOPSC item-10: from the Hospital Survey on Patient Safety Culture. "Hospital units work well together to provide the best care for patients".*

**Supplementary table 7**: Bivariate analysis (linear regressions) of variables at healthcare workers level on fatigue level (Pichot score) at inclusion. Variables were sorted according to their p-values. Variables in bold (p_value > 0 .20) were not considered in the multivariate analysis, expected the "commuting time to work" variable.

| **Variables** | **p_value** |
|---|---|
| Work overcommitment (Siegrist score) | 5.86 e-38 |
| Presence at work while sick | 2.07 e-14 |
| HSOPSC item-1[a] | 3.09 e-12 |
| HSOPSC item-8[b] | 2.71 e-10 |
| HSOPSC item-10[d] | 1.41 e-07 |
| Professional life events | 1.12 e-06 |
| Support from hierarchy (Karasek score) | 2.09 e-06 |
| Gender | 1.49 e-05 |
| Age | 0.0002 |
| 48 hours weekly rest | 0.0002 |
| Breaks cancelled | 0.0003 |
| Personal life events | 0.0008 |
| HSOPSC item-9[c] | 0.0016 |
| Participation to the schedule | 0.0036 |
| Irregularity of meal time | 0.0120 |
| Number of breaks per day | 0.0128 |
| Work schedule variation | 0.0172 |
| Years of work in the hospital | 0.0239 |
| Number of visits to the Occupational safety and health (OSH) department | 0.0377 |
| Overtime hours | 0.0506 |
| Knowledge of the schedule | 0.0810 |
| Professional status | 0.0925 |
| Support from colleagues (Karasek score) | 0.1123 |
| Number of children (between 0 and 10 years old) | 0.1665 |
| Nightshift/duty on last months | 0.1679 |
| **Schedule assignment frequency** | 0.2138 |
| Commuting time to work | 0.2360 |
| **Marital status** | 0.5132 |
| **Transportation** | 0.5841 |
| **Supervising responsibility** | 0.6387 |
| **Work schedule of last months** | 0.6860 |
| **Profession** | 0.7095 |
| **Total duration of breaks per day (min)** | 0.8452 |
| **Weekly working hours** | 0.9533 |
| **Number of nightshift/duties** | 0.9789 |

[a]*HSOPSC item-1: from the Hospital Survey on Patient Safety Culture. "Hospital management provides a work climate that promotes patient safety".*

[b]*HSOPSC item-8: from the Hospital Survey on Patient Safety Culture. "The actions of hospital management show that patient safety is a top priority".*

[c]*HSOPSC item-9: from the Hospital Survey on Patient Safety Culture. "Hospital management seems interested in patient safety only after an adverse event happens".*

[d]*HSOPSC item-10: from the Hospital Survey on Patient Safety Culture. "Hospital units work well together to provide the best care for patients".*
